# Apolipoprotein A1 levels and its association with NS1 in the pathogenesis of acute dengue

**DOI:** 10.1101/2022.07.20.22277834

**Authors:** Deshni Jayathilaka, Laksiri Gomes, Chandima Jeewandara, Dhanushka Herath, Geethal. S. Bandara Jayarathna, Ananda Wijewickrama, Graham S. Ogg, Gathsaurie Neelika Malavige

## Abstract

**Background:** The dengue NS1 antigen is a secretory protein and was shown to associate with apolipoprotein A1 (ApoA1). Therefore, we sought to investigate the changes in ApoA1 levels in patients with varying severity of acute dengue and then proceeded to investigate how ApoA1 and NS1 interactions affect cytokine production from primary human monocytes.

**Methodology/Principal Findings:** Serial measurements of ApoA1, viral loads, NS1 antigen levels and lipid profiles were done in adult patients with dengue fever (DF= 21) and dengue haemorrhagic fever (DHF =28). To investigate the effect of ApoA1-NS1 interactions in cytokine production from primary human monocytes, cells were harvested from healthy individuals (n=6) and co-cultured with varying concentrations of ApoA1 and levels of IL-6 and TNF-α measured in culture supernatants.

The ApoA1 levels were significantly lower in patients with DHF on day 5 of illness (p=0.04) compared to those with DF. ApoA1 levels did not show any correlation with either the NS1 antigen levels or the viral loads in patients with DF or DHF, although they did significantly and inversely correlate with liver transaminases, AST (Spearmans R=-0.55, p <0.0001) and ALT (Spearmans R=-0.49, p<0.0001) in patients with DF. However, a significant correlation was not seen between ApoA1 levels and AST levels (Spearmans R=-0.45, p=0.09) in patients with DHF and ALT levels (Spearman’s R=0.0.5, p=0.64). HDL and LDL levels were significantly lower on day 5 of the illness in patients with DHF compared to those with DF. Co-culture of NS1 and ApoA1 resulted in an increased IL-6 production (but not TNFα) in culture supernatants in a dose dependent manner.

**Conclusions/Significance:** HDL, LDL and ApoA1 levels were significantly lower in those who have severe dengue, especially in the critical phase and ApoA1 levels inversely correlated with the extent of rise in liver enzymes. While co-culture of ApoA1 with NS1 in primary human monocytes induced high IL-6 levels in a dose dependent manner, this was not seen for TNFα suggesting that the interaction of ApoA1 with NS1 could give rise to different outcomes.

**Author summary:** The dengue NS1 antigen is a secretory protein and was shown to associate with apolipoprotein A1 (ApoA1). Therefore, we sought to investigate the changes in ApoA1 levels in patients with varying severity of acute dengue and then proceeded to investigate how ApoA1 and NS1 interactions affect cytokine production from primary human monocytes. Serial measurements of ApoA1, viral loads, NS1 antigen levels and lipid profiles were done in adult patients with dengue fever (DF) and dengue haemorrhagic fever (DHF). HDL, LDL and ApoA1 levels were significantly lower in those who have severe dengue, especially in the critical phase and ApoA1 levels inversely correlated with the extent of rise in liver enzymes. While co-culture of ApoA1 with NS1 in primary human monocytes induced high IL-6 levels in a dose dependent manner, this was not seen for TNFα, suggesting that the interaction of ApoA1 with NS1 could give rise to different outcomes.

## Introduction

Infection due to the dengue virus (DENV) was named as one of the top ten threats to global health by the WHO in 2019 [1]. 390 million individuals are estimated to be infected with the DENV each year [2], with the highest incidence, deaths and age-standardized disability-adjusted life years (DALYs) rates being highest in South Asia and South East Asia [3]. Although the majority of infections manifest as asymptomatic infection, undifferentiated fever or dengue fever, some develop dengue haemorrhagic fever (DHF)/ dengue shock syndrome (DSS) or dengue infection complicated with organ failure [4]. Unfortunately, there is no specific treatment for this infection nor a licensed safe and effective vaccine, which is effective against all four serotypes of the virus.

The DENV comprises of 3 structural and 7 nonstructural proteins [5]. DENV NS1 protein is one of the structural proteins and is a ∼48kDa glycoprotein that is absent from the virion but exists as either as a dimeric membrane associated form or as a tetramer or hexamer in the secretory (sNS1) form [6, 7]. The secreted form of NS1 by itself has shown to result in disease pathogenesis by disrupting the endothelial glycocalyx [8, 9], cytokine production from immune cells [10], inducing inflammatory phospholipase A2 enzymes [11] and immune evasion by interfering with complement activation [12]. Although some studies have shown that high NS1 levels associate with increase in disease severity[13, 14], some have not found such association [15]. Further, it has been shown that the NS1 antigen levels vary according to the DENV serotype and immune status [16, 17]. The NS1 in certain DENV2 strains such as the D2Y98P DENV2, was not shown to cause disease pathogenesis or vascular leak in certain mouse models, although this strain did cause lethal infection in mice [18]. Pathogenesis of NS1 was shown to be strain dependent and the level of NS1 antigenaemia in acute illness, due to the same DENV serotype dependent on certain critical mutations within NS1, increasing its virulence [19, 20]. Therefore, many factors appear to determine the virulence of NS1 antigen in dengue infection.

The secreted hexameric NS1 forms an atypical barrel structure similar to the smallest subclass of high-density lipoproteins (HDL)[21]. This hexameric NS1 has shown to associate with many types of such as triglycerides, cholesterol esters, phospholipids and sphingomyelin [21]. It was recently shown that the lipid composition of NS1 hexamer was different in NS1 antigens associated with increased virulence, compared to those who were associated with milder disease [20]. NS1 complex in the supernatants of AD293 transfected cell lines, was shown to coimmunoprecipitate with apolipoprotein 1 (ApoA1)[22] and NS1 antigen was shown to directly interact with ApoA1 in patients [23, 24]. Interaction of ApoA1 opposes the action of NS1 on lipid rafts in cells and thereby reducing the infectivity of the DENV [23]. In contrast, in a different study ApoA1 increased the infectivity of dengue viral particles by facilitating entry of the virus into the cells through SR-BI, which is the major cell receptor for HDL[25]. A recent study showed that patients with severe dengue had higher ApoA1 levels, in samples collected from patients during day 0 to 10 since onset of symptoms [26]. However, it has been shown in many studies, that the levels of different inflammatory mediators and other biochemical markers vary widely based on the day and time of the day of obtaining samples from patients [27-29]. Therefore, we initially sought to investigate the changes in ApoA1 levels in patients with varying severity of acute dengue and then proceeded to investigate how ApoA1 and NS1 interactions affect IL-6 and TNF-α production from primary human monocytes.

## Methods

### Patients and healthy participants

Forty nine adult patients with varying severity of acute dengue infection were recruited from the National Institute of Infectious Disease, Sri Lanka following informed written consent. Daily consecutive blood samples were obtained from patients from the day of admission to the hospital until they were discharge from the hospital, capturing the course of illness. The day on which the patient first developed fever was considered day one of illness. Only those whose duration from onset of illness was ≤4 days were recruited. Patient were followed up throughout their hospital stay and clinical features such as fever, blood pressure, pulse pressure and the urine output were measured at least 4 times a day. Serial recordings of liver function tests, viral loads, full blood counts and the assessment of the extent of fluid leakage were done daily. Fluid leakage was assessed by ultrasound scans to detect fluid in pleural and peritoneal cavities. Clinical disease severity was classified according to the 2011 World Health Organization (WHO) dengue diagnostic criteria[4]. Patients with a rise in haematocrit ≥ 20% of baseline or with clinical or ultrasound scan evidence of plasma leakage were classified as having DHF. The presence of cold clammy skin, along with a narrowing of pulse pressure of ≤20 mmHg was defined as a shock. Accordingly, 21 patients were classified as DF and 28 patients were classified as DHF (Table 1).

**Table 1.**
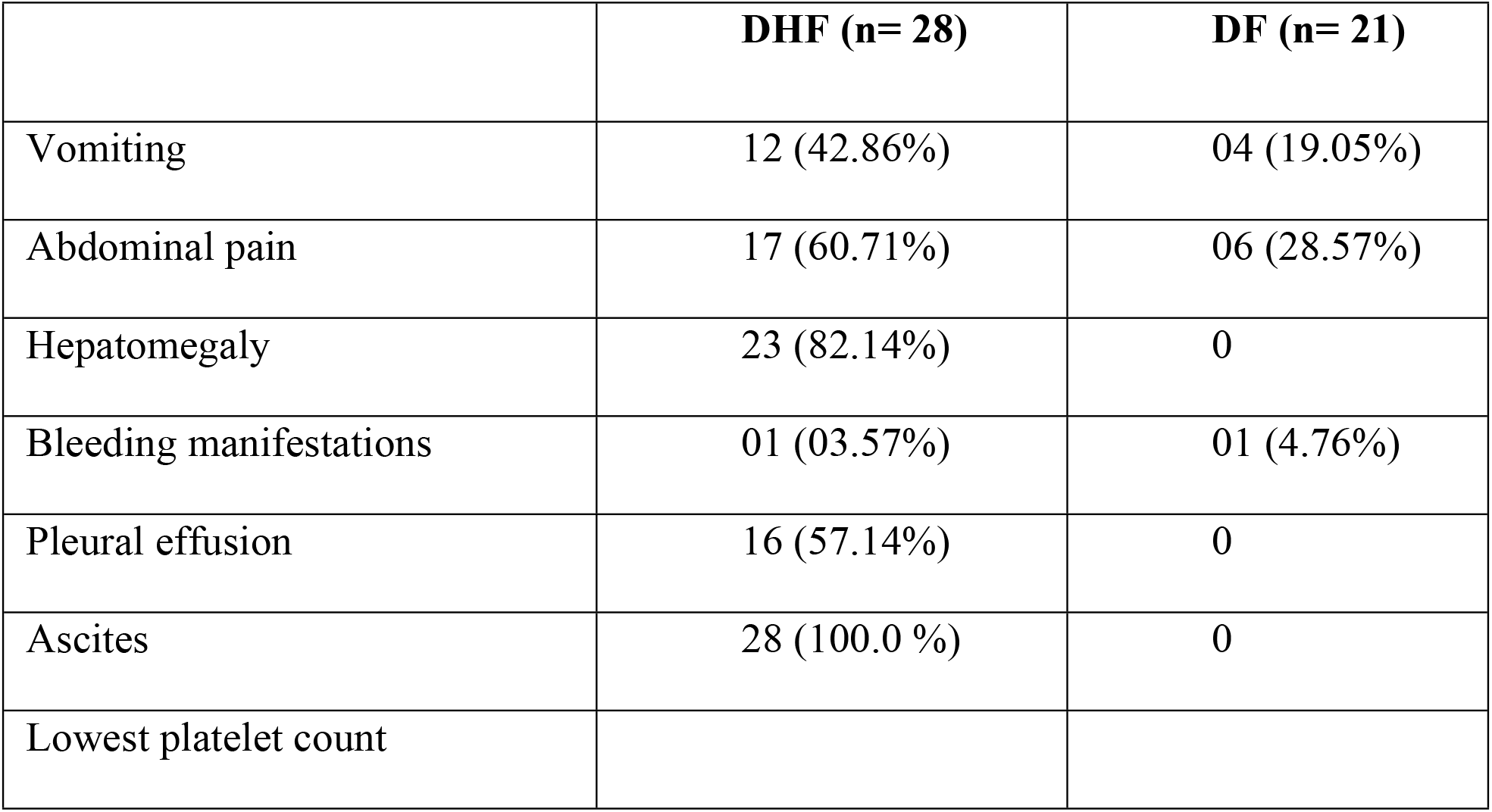
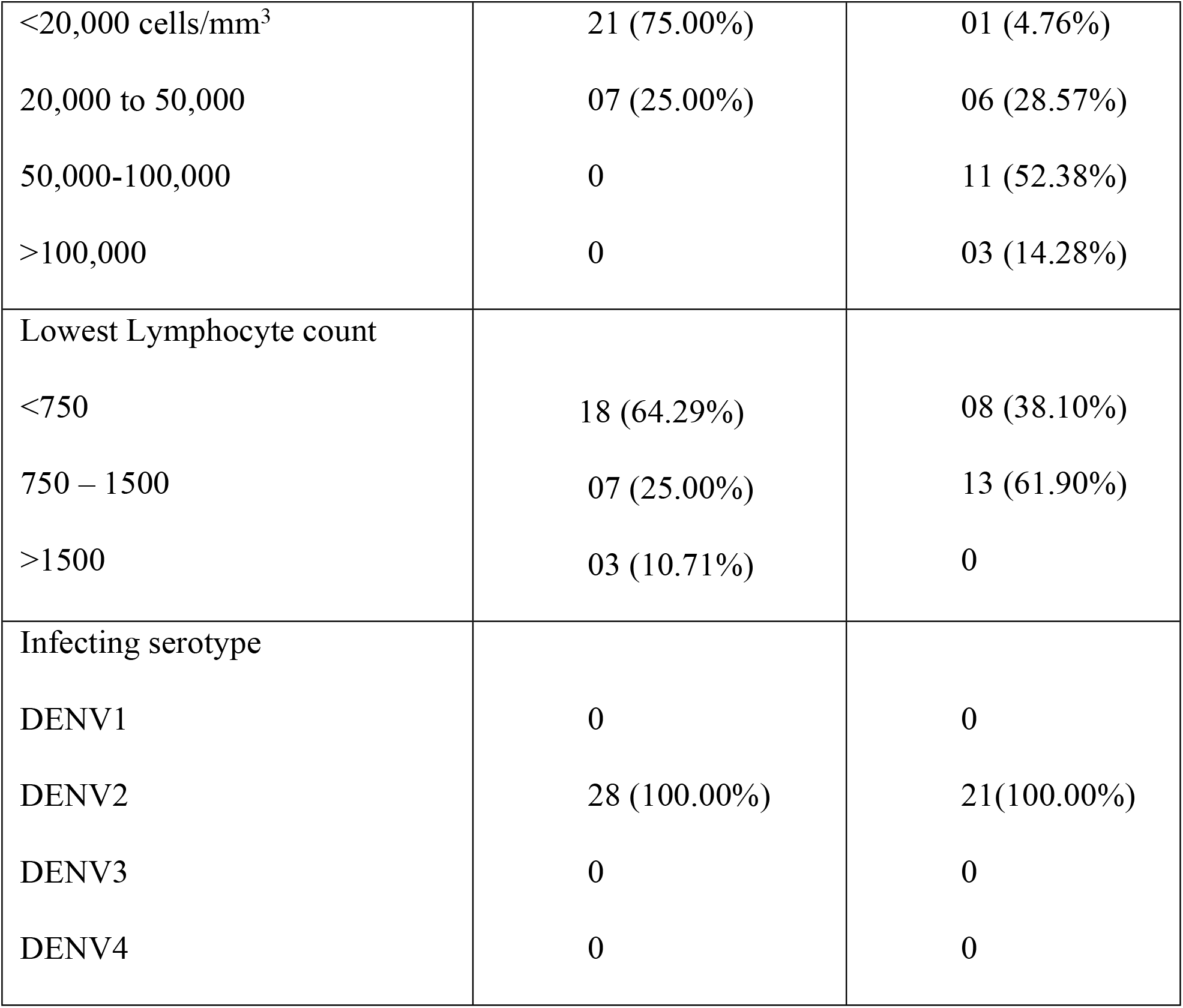
Clinical and laboratory characteristics of acute dengue patients with DHF and DF recruited for the study.

|  | **DHF (n= 28)** | **DF (n= 21)** |
| --- | --- | --- |
| Vomiting | 12 (42.86%) | 04 (19.05%) |
| Abdominal pain | 17 (60.71%) | 06 (28.57%) |
| Hepatomegaly | 23 (82.14%) | 0 |
| Bleeding manifestations | 01 (03.57%) | 01 (4.76%) |
| Pleural effusion | 16 (57.14%) | 0 |
| Ascites | 28 (100.0 %) | 0 |
| Lowest platelet count |  |  |
| <20,000 cells/mm<sup>3</sup> | 21 (75.00%) | 01 (4.76%) |
| 20,000 to 50,000 | 07 (25.00%) | 06 (28.57%) |
| 50,000-100,000 | 0 | 11 (52.38%) |
| >100,000 | 0 | 03 (14.28%) |
| Lowest Lymphocyte count |  |  |
| <750 | 18 (64.29%) | 08 (38.10%) |
| 750 – 1500 | 07 (25.00%) | 13 (61.90%) |
| >1500 | 03 (10.71%) | 0 |
| Infecting serotype |  |  |
| DENV1 | 0 | 0 |
| DENV2 | 28 (100.00%) | 21(100.00%) |
| DENV3 | 0 | 0 |
| DENV4 | 0 | 0 |

### Ethics statement

Fourteen healthy individuals, with a normal lipid profile were recruited from the University of Sri Jayewardenepura to determine ApoA1 levels in the healthy population. Ethical approval was obtained by the Ethics Review Committee of the Faculty of Medical Sciences, University of Sri Jayewardenepura. All patients and healthy individuals were recruited following informed written consent.

### Determining viral loads and serotype of the DENV

Acute dengue infection was confirmed by detection of virus by quantitative real time PCR and DENV viruses were serotyped, and viral copy numbers were determined as previously described [30]. Briefly, RNA was extracted from serum samples using QIAamp Viral RNA Mini Kit (Qiagen, USA) according to the manufacturer’s protocol. Multiplex quantitative real-time PCR was performed as previously described using the CDC real time PCR assay for detection of the DENV and modified to quantify DENV. Oligonucleotide primers and a dual labeled probe for DEN 1,2,3,4 serotypes were used (Life technologies, India) based on published sequences. To quantify viruses, standard curves of DENV serotypes were generated as previously described in Fernando, S. *et.al* [30].

### Assays for detection of NS1 antigen and dengue IgM and IgG antibodies

The presence of NS1 antigen was assessed using the NS1 early dengue enzyme-linked immunosorbent assay (ELISA) (Panbio, Brisbane, QLD, Australia) according to the manufacturer’s instructions. NS1 antigen levels were semi quantitatively assessed in serial blood samples of patients and expressed as Panbio units. Dengue antibody assays were performed using a commercial capture-IgM and IgG ELISA (Panbio, Brisbane, Australia) [31]. Based on the WHO criteria, patients with an IgM: IgG ratio of >1.2 were considered to have a primary dengue infection, while patients with IgM: IgG ratios <1.2 were categorized under secondary dengue infection [32].

### Quantitative cytokine assays

Quantitative ELISA for Apolipoprotein A1 (Mabtech, Sweden) was performed on patient and serum samples from healthy individuals and quantitative ELISA for IL-6 (Mabtech, Sweden) and TNF-α (Biolegend, USA) were performed on primary monocyte culture supernatants according to the manufacturer’s instructions.

### Assessing the lipid profile

Lipid profiles (HDL, total cholesterol, LDL, VLDL and triglyceride levels) were measured using Thermo Scientific™ system reagents with Indiko™ (Thermo Fisher Scientific, Germany) fully automated chemistry analyzer. The analysis were performed using standard settings as per the manufacturer recommendation. Due to the limited availability of serum from the above cohort of patients, changes in HDL, LDL, VLDL, total cholesterol and triglyceride levels were measured in a subset of the above cohort of patients. Therefore, the lipid profiles were only done in 15/22 patients with DF and 16/27 patients with DHF.

### Determining the effect of ApoA1 and NS1 in potentiating cytokine production

Peripheral Blood Mononuclear cells (PBMCs) were obtained from 6 volunteers using lymphoprep (Axis shield) density gradient centrifugation. The monocytes were positively selected from PBMCs using CD14 magnetic beads (Milteny Biotech, Germany) and MACS separation columns (Milteny Biotech, Germany). The monocyte purity was determined by flowcytometry and was between 90-95%. Isolated primary human monocytes (1×10^5^ cells) were co-cultured with 1µg/ml of dengue NS1 recombinant protein (Native antigen, UK) and varying concentrations of Apolipoprotein A1(Abcam, UK) for 16 hours. The monocytes were cultured in RPMI (Life technologies, USA) supplemented with 10% AB negative human serum (Sigma Aldrich, USA), 1% Penicillin / Streptomycin (Sigma, USA) and 1% L-glutamine and was dispensed in a volume of 200µl per well, in 96 well round bottom plates (Corning, USA). Human Apo-A1 was used at concentrations of 1.68µg/ml, 10µg/ml, 65µg/ml, 100µg/ml and 195µg/ml to co-culture with monocytes alone and together with 1µg/ml of dengue NS1 recombinant protein. All experiments were carried out in duplicates. The supernatant was harvested after 16 hours of co-culture of Apo-AI and NS1 at various concentrations at 37°C supplemented with 5% CO2 and was then used to determine IL-6 and TNFα levels.

### Statistical analysis

Statistical analysis was performed using GraphPad Prism version 8. As the data were not normally distributed (as determined by the frequency distribution analysis of GraphPad Prism), non-parametric tests were used in the statistical analysis and two-sided tests were carried out in all instances. Spearman rank order correlation coefficient was used to evaluate the correlation between serum ApoA1 levels, NS1 antigen levels, viral loads and liver transaminases. Differences in the serial values of ApoA1 and NS1 antigen in patients with DHF and DF and differences in IL-6 and TNF-α levels in monocyte culture supernatants were determined using the Holm-Sidak method. Corrections for multiple comparisons were done and the statistically significant value was set at 0.05 (alpha). Differences in mean values of Apo-A1 between healthy individuals and individuals with DF and DHF were compared using the Mann-Whitney U test (two tailed).

## Results

### Kinetics of Apolipoprotein A1 in patients with acute dengue infection

Based on WHO 2011 guidelines, 21 patients were classified as DF and 28 patients were classified as DHF (table 1). None of the patients developed shock. The ApoA1 levels were significantly lower in patients with DHF (n=28) on day 5 of illness (p=0.04) compared to those with DF (n=21) (Fig 1A), when all the patients were analyzed together, irrespective of gender. However, as the ApoA1 levels are different among men and women with women having higher levels, we also analysed the changes in ApoA1 based on gender [33]. Although, the ApoA1 levels were lower in both male and female patients with DHF, this was not significant, probably due to the lower number of female patients with DF (Fig 1B and 1C). However, despite lower number of females in the DF group (6/22), when analyzing the overall changes of ApoA1, the levels were still significantly lower in patients with DHF.

**Figure 1:**
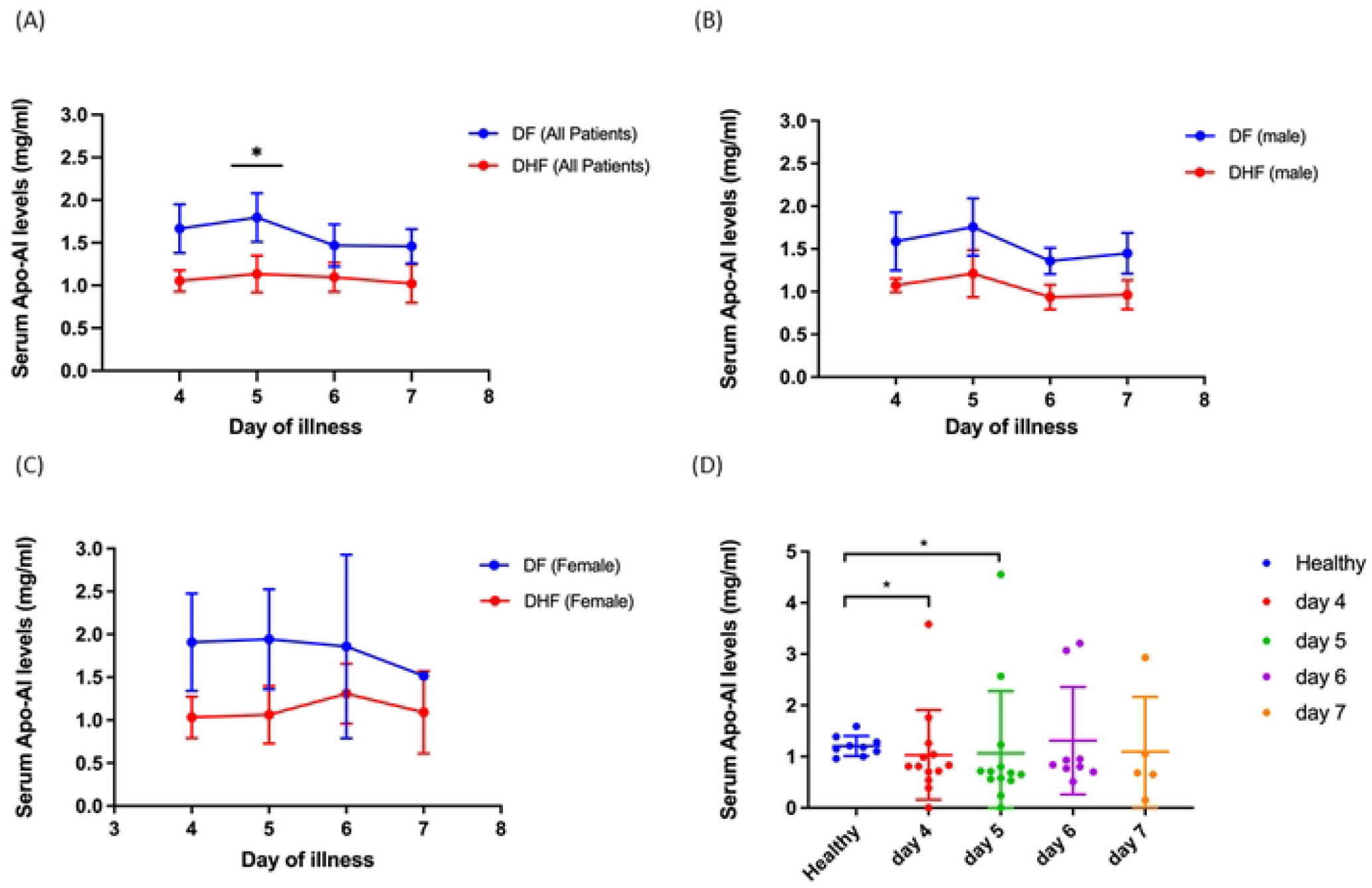
Kinetics of ApoAI levels in acute dengue. (a) ApoA1 levels were measured in patients with DHF (n=28) and DF (n=21) (b) in male patients with DF (n=15) and DHF (n=14) (c) in female patients with DF (n=6) and DHF (n=14) using a quantitative ELISA. ApoA1 levels in patients with DHF are indicated in red, and in those with DF in blue. The ApoA1 [levels were also compared in female patients with DHF (n=14) and healthy women (n=9) (D). The Holm-Sidak method was used to compare the levels between patients with DF and DHF at different days and the Mann-Whitney test was used to compare the levels between females with DHF and healthy females. All tests were two tailed. Error bars in A to C indicate mean and standard error of mean (SEM). Error bars in D indicate the median and the interquartile range. *p<0.05

We also measured the ApoA1 levels in healthy individuals (those who were not obese, dyslipidaemic and did not have diabetes or hypertension). The ApoA1 levels of females (median = 1.18; interquartile range (IQR) = 1.05 – 1.34) were slightly higher than males (median = 1.08; interquartile range (IQR) = 0.97 – 1.23). Furthermore, females with DHF at day 4 (p=0.035) and day 5 (p=0.02) had significantly lower ApoA1 levels compared to healthy female individuals (Fig 1D).

### Association of Apo AI with NS1 antigen levels, viraemia and laboratory parameters

In order to determine the relationship between serum ApoA1 levels and NS1 antigen, we semi-quantitatively measured dengue NS1 antigen levels by using the Panbio NS1 capture ELISA throughout the course of illness. The serum ApoA1 levels did not show any correlation with either the NS1 antigen levels of the viral loads in patients with DF or DHF. ApoA1 levels did significantly inversely correlate with liver transaminases, AST (Spearmans R=-0.55, p <0.0001) and ALT (Spearmans R=-0.49, p<0.0001), in patients with DF, probably due to lower production. However, a significant correlation was not seen between ApoA1 levels and AST levels (Spearmans R=0.016, p=0.9) in patients with DHF (Fig 2A) and ALT levels (Spearman’s R=0.05, p=0.64) (Fig 2B).

**Figure 2:**
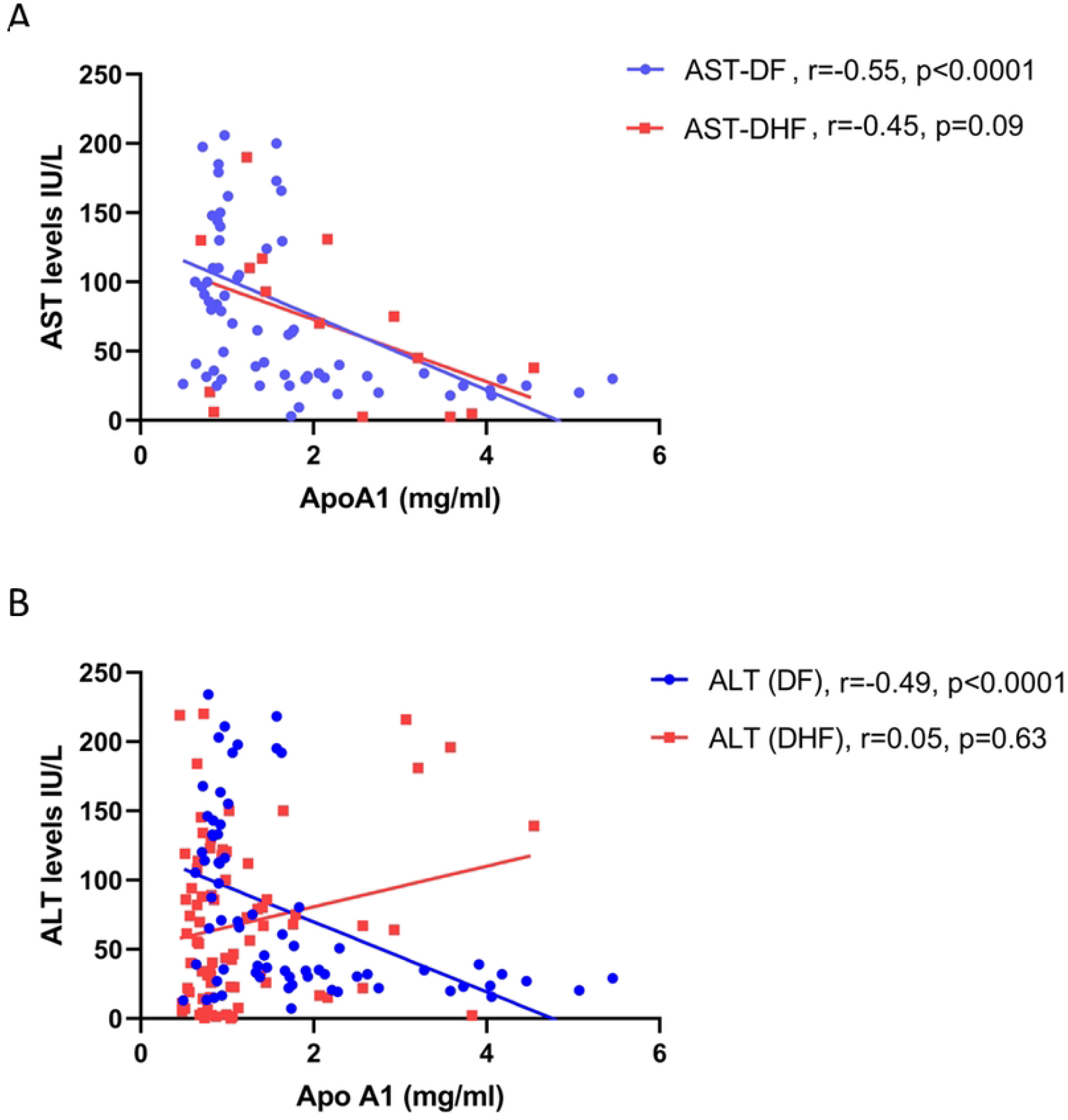
Association of ApoA1 levels with liver transaminases. Serum ApoA1 levels were correlated with patients with DF (n=21) and DHF (n=28), during the time course of illness with serum AST levels (A) and ALT levels (B). Spearman rank order correlation coefficient was used to evaluate the correlation between serum ApoA1 levels and liver transaminases.

### Changes in lipid profiles throughout the course of illness in patients with acute dengue

As we observed changes in ApoA1 levels throughout the course of illness, we then sought to assess the changes in the lipid profiles in these changes. Due to the limited availability of serum, we measured the lipid profiles of 15/22 patients with DF and 16/27 patients with DHF. The HDL levels were significantly lower on day 4, 5 and 6 of the illness in patients with DHF compared to those with DF (Fig 3A). LDL levels were also significantly lower in patients with DHF, compared to those with DF on day 5 of the illness, although the difference was not as marked as with HDL levels (Fig 3B). The levels of VLDL and triglyceride were not significantly lower in patients with DHF compared to patients with DF, although these levels too remained low, throughout the course of illness (Fig 3C and 3D). The total serum cholesterol levels were also significantly lower in those with DHF compared to those with DF on day 5 and 6 of illness (Fig 3E).

**Figure 3:**
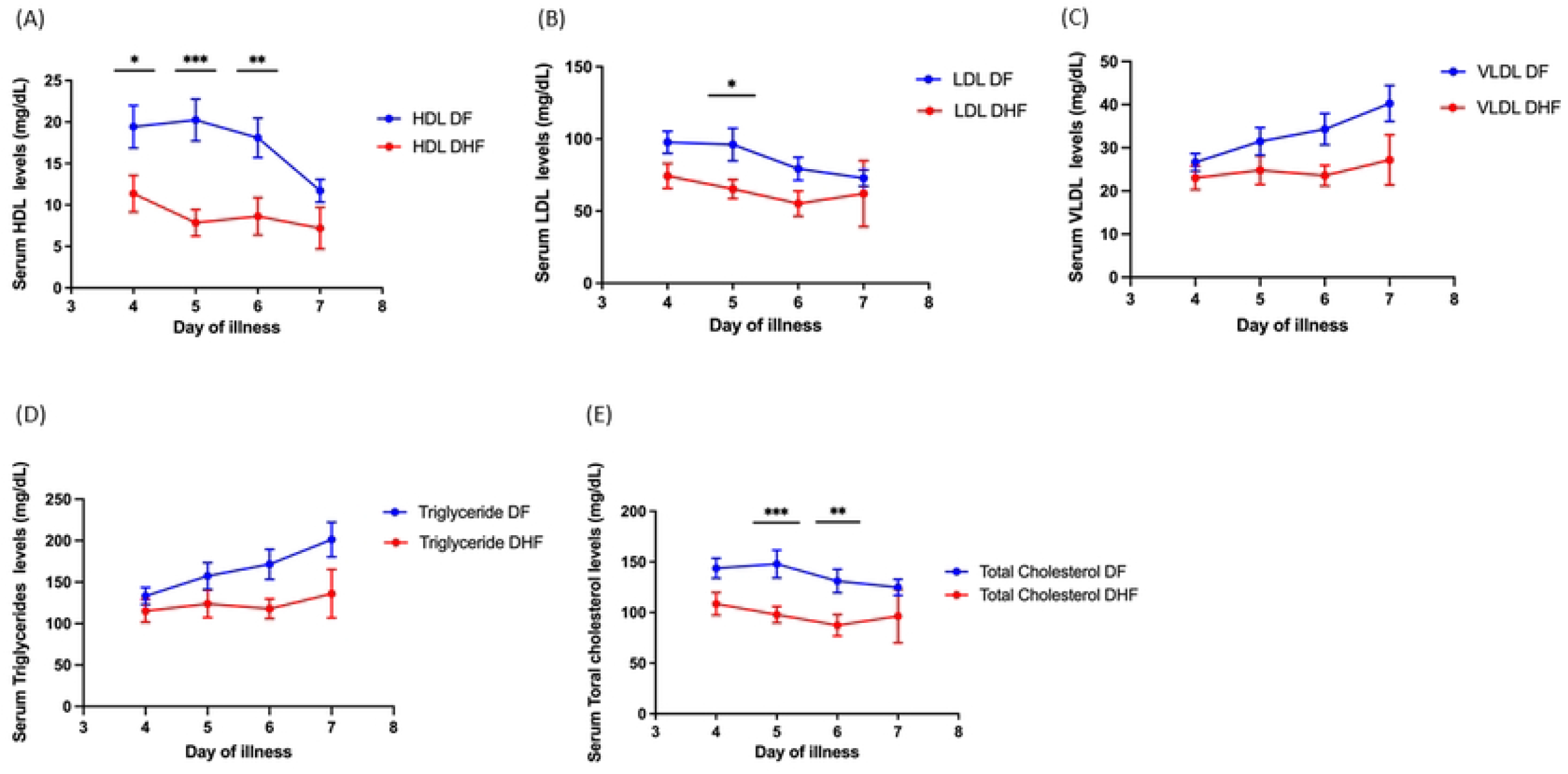
Kinetics of serum cholesterol levels in acute dengue. (a) HDL levels (b) LDL levels (c) VLDL levels (d) triglyceride levels and (e) total cholesterol levels were measured throughout the course of illness in patients with DHF (n=15) and DF (n=16). The cholesterol levels in patients with DHF are indicated in red, and in those with DF in blue. Error bars indicate mean and standard error of mean (SEM). *P<0.05, **P<0.01, *** P<0.001

### Effect of ApoA1 and NS1 on primary human monocytes in inducing inflammatory cytokines

As there were marked differences between ApoA1 and cholesterol subfractions between patients with DF and DHF, and since it was shown that ApoA1 binds to DENV NS1[24], we wished to evaluate the possible effects of ApoA1 and NS1 together on primary human monocytes. In order to determine the effect of ApoA1 along with NS1, primary monocytes of 6 individuals were co-cultured with varying concentrations of natural human ApoA1 (specified by the manufacturer as endotoxin free) at various concentrations (195μg/ml, 100μg/ml, 65μg/ml, 10μg/ml, and 1.68μg/ml) together with DENV1 NS1 antigen, that was specified by the manufacturer as endotoxin free (1μg/ml) (Native antigen, UK) for 16 hours. IL-6 and TNF-α levels were assessed in culture supernatants by quantitative ELISA. Culture of the monocytes with NS1 alone resulted in IL-6 production by the monocytes as described earlier [10, 11] with median value of 1,197 pg/ml (IQR 176.5 to 2465 pg/ml). However, co-culture of NS1 and ApoA1 resulted in an increased IL-6 production in culture supernatants in a dose dependent manner (Fig 4A). At ApoA1 concentrations of 10μg/ml when co-cultured with NS1 there was a significant difference of IL-6 production compared to NS1 alone. For instance, the median IL-6 production by monocytes was 3,492 pg/ml (IQR 2,778 to 4,633 pg/ml) whereas it was 1638 pg/ml (IQR 904 to 3317 pg/ml) for Apo A1 alone. However, at higher ApoA1 concentrations, ApoA1 alone appeared to induce production of IL-6 (Fig 4B).

**Figure 4:**
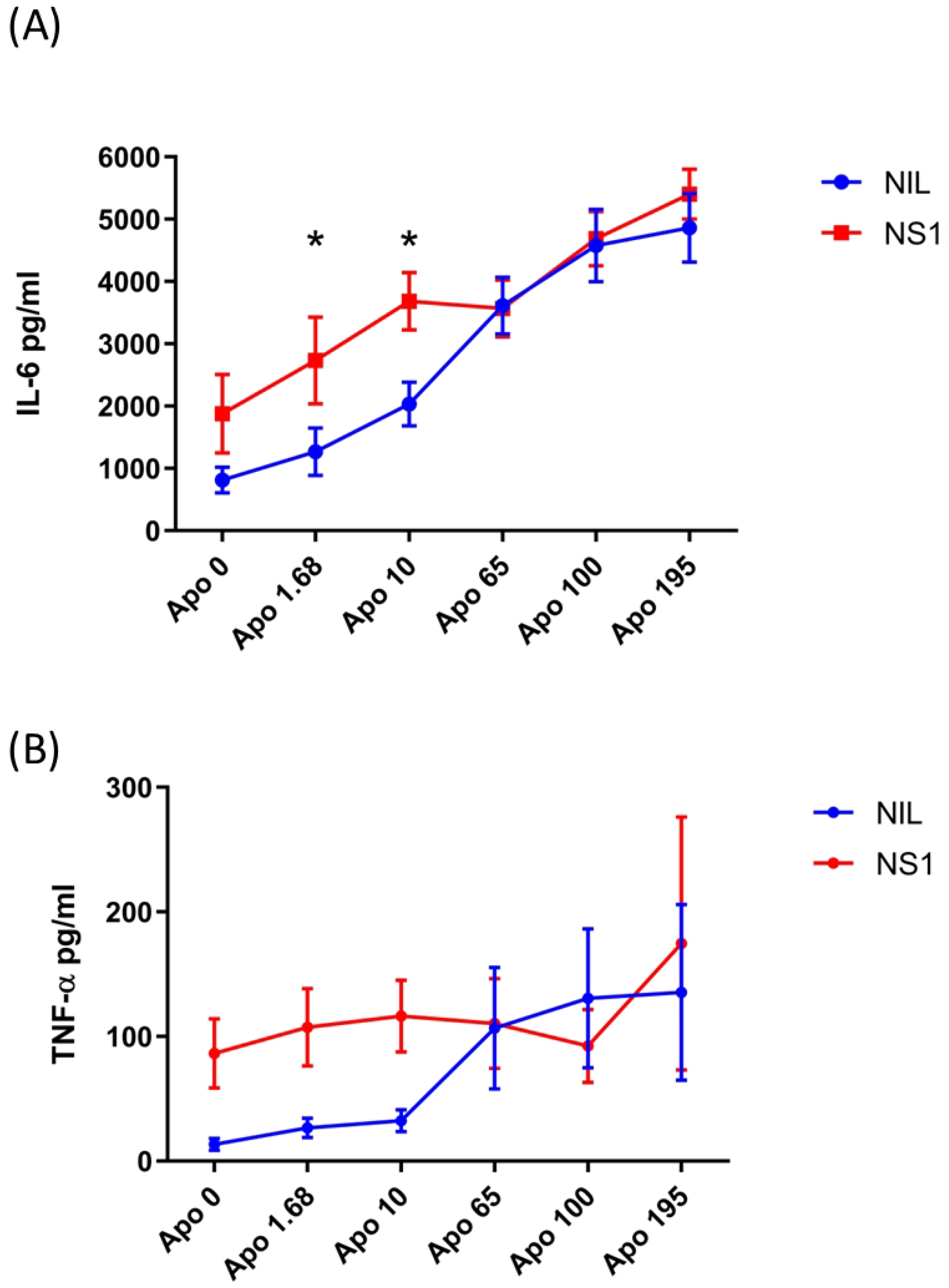
Monocyte responses to co-culture of NS1 and Apo-AI. Varying concentration of ApoA1 was co-cultured with 1µg/ml of DENV1 NS1 with primary human monocytes of 6 individuals for 16 hours and IL-6 (A) and TNFα (B) was measured in the monocyte culture supernatants by ELISA. The Holm-Sidak method was used to compare the levels between different levels of ApoA1 co-cultured with NS1 or alone. All tests were two tailed. Error bars indicate mean and standard error of mean (SEM). *p<0.05

NS1 alone caused significant production of TNFα by the monocytes (median 64.7 pg.ml, IQR). However, co-culture of NS1 with ApoA1 did not appear to increase the TNFα levels substantially, when compared to NS1 alone (Fig 3C). However, as observed with IL-6, Apo-AI concentrations above 65µg/ml induced TNFα production by monocytes by itself.

## Discussion

In this study we have shown that ApoA1 levels are lower in patients with DHF compared to those with milder forms of dengue. The levels were significantly lower especially in female patients with DHF, compared to healthy females. Along with ApoA1, serum HDL and total cholesterol levels were lower with no changes in serum triglycerides and VLDL levels. Previous studies, which had looked at one time point, rather than the changes throughout the course of illness had shown that serum cholesterol and LDL levels, but not HDL levels were lower in those who developed severe dengue [34], while some showed that HDL levels were also low [35]. Non-survivors of sepsis were found to have lower serum cholesterol, HDL and ApoA1; and ApoA1 alone was shown to correlate with 30-day mortality in sepsis [36]. HDL and ApoA1 were also found to be a marker of liver disease and low levels were seen in patients with liver disease due to reduced synthesis [37]. We too observed that AST levels and ALT levels (in dengue fever) inversely correlated with the extent of rise in liver enzymes, suggesting that liver dysfunction, which is common in dengue [30], could have resulted in lower levels.

Although the exact mechanisms leading to lower HDL in dengue is not known, cytokines such as TNFα and IL-1β have shown to associate with reduction in the activity of lecithin cholesteryl acyl transferase, resulting in a lower levels of cholesterol esters available for generation of HDL and LDL[35]. In addition, protein and lipid content of HDL is shown to be altered in inflammation leading to a reduction of ApoA1 and replacement of ApoA1 with inflammatory proteins such as complement [38]. Two of the components of HDL, sphingosine-1-phosphate (S1P) [39] and ApoA1 were shown to be significantly lower in patients with severe disease, especially in the critical phase. Both of these have been shown to be important in maintaining the endothelial barrier function [38, 39]. In addition, ApoA1 was shown to bind to LPS resulting in reduction in the generation of proinflammatory cytokines by LPS [38]. High serum LPS levels were seen in patients with severe dengue and LPS was shown to act synergistically with the DENV to induce production of many proinflammatory cytokines from monocytes [40, 41]. Therefore, the reduction in ApoA1 available for binding of LPS, leading to inhibiting its effects, seem to be impaired in patients with severe dengue due to the lower levels of ApoA1 available.

However, in dengue, ApoA1 appears to have a dual role. Although ApoA1 could be having a protective role by binding to LPS and by inducing many other anti-inflammatory effects, its interaction with NS1 appears to lead to disease pathogenesis. We found that while ApoA1-NS1 interactions increased IL-6 production from primary human monocytes, a significant increase in TNFα was not seen. Therefore, while NS1-ApoA1 interaction may induce proinflammatory cytokines, the type and quantity of cytokines and other inflammatory mediators may depend on the composition of Apo-A1 within NS1 and other lipid cargo included in the NS1 hexamer.

In conclusion, we have shown that serum cholesterol, HDL and ApoA1 were significantly lower in those who have severe dengue, especially in the critical phase and ApoA1 levels inversely correlated with the extent of rise in liver enzymes. While co-culture of ApoA1 with NS1 in primary human monocytes induced high IL-6 levels in a dose dependent manner, this was not seen for TNFα suggesting that the interaction of ApoA1 with NS1 could give rise to different outcomes.

## Data Availability

All relevant data are within the manuscript and its Supporting Information files.

## Acknowledgements

We are grateful to the Centre for Dengue Research, and the UK Medical Research Council for funding.

